# Fall incidence and risk among adults with cerebral palsy

**DOI:** 10.64898/2025.12.17.25342293

**Authors:** Lauren N. Kang, Linda E. Krach, Elizabeth R. Boyer

## Abstract

**Purpose:** Falls are common in adults with cerebral palsy (CP) and remain a significant concern. The study examined whether fall risk, fall incidence, and fall-related injuries differ by GMFCS level, CP type, age, and gender in adults with CP.

**Materials and Methods:** A cross-sectional, retrospective chart review was conducted for adults 18 years or older with CP who received outpatient specialty care between December 2022 to May 2023. CP diagnosis and type were determined using ICD-10 codes; GMFCS level was extracted from electronic medical record notes. Fall risk was determined using the Morse Fall Scale (MFS), with scores ≥45 indicating high fall risk.

**Results:** Among 647 adults with CP, ambulatory individuals (GMFCS II–III) had significantly higher MFS scores and fall incidence than non-ambulatory individuals (p < 0.001). CP type and gender showed significant group differences, whereas age did not. Individuals with spastic diplegia and unspecified CP showed higher fall risk. Females were twice as likely as males to report a recent fall, although MFS scores did not differ by gender. Five injuries were reported among ambulatory individuals in the past three months.

**Conclusions:** Findings highlight the importance of refining fall-risk assessment and prevention strategies for higher-risk groups.

## Introduction

Cerebral palsy (CP) is a condition caused by a nonprogressive perinatal brain injury or atypical brain development that affects sensorimotor functions, primarily impacting individuals’ ability to balance and move, though other body systems can be affected.^1,2^ Owing to these impairments, falls are common throughout life,^3–7^ as opposed to the general population where falls are most common in the first few years of life and then later as an older adult. Accumulating evidence in the CP literature illustrates that falls cause physical injuries and negative emotions like embarrassment, shame, or frustration, all of which can affect independence, confidence, self-esteem, physical activity, participation, and quality of life.^3,5–8^ Due to adults’ concern about falling, some individuals avoid daily activities as reported on the Short Falls Efficacy-International Avoidance Behavior survey,^3^ and Bell et al. found that 12-33% chose not to leave their home and avoid exercise or recreational/leisure activities.^9^ Such decisions may have immediate and long-term repercussions on physical and mental health, highlighting the urgency to address falls and concerns about falling. Adults with CP report that falls and their repercussions have greater effects on their wellbeing in adulthood than childhood.^10,6^ Identifying adults’ risk of falls and injuries (i.e., fall risk stratification) is the first step toward addressing this lifelong concern, and it is the first step in the World Falls Guidelines framework for fall prevention and management in older adults.^11^

Understanding patient factors that contribute to falls and injury risk can help increase awareness in patients and inform clinical interventions or behavior modification; however, fall risk factors for individuals with CP have not been robustly studied. Previous studies have shown that falls are more common among individuals with CP who can ambulate compared to those who cannot,^4^ and among the subgroup that ambulates, those with moderate level of impairment but who do not use a walking device (i.e., Gross Motor Function Classification (GMFCS) level II) fall more often that those with less impairment or those with greater impairment but who regularly use walking devices (i.e., GMFCS level III).^4,3,12^ It remains unknown whether individuals who have hemiplegic/unilateral involvement are at decreased risk of falls or injuries than individuals who have both legs affected by their CP. In the general adult population, fall incidence increases from young to middle to older adulthood.^13,14^ The limited evidence in predominantly young- to middle-aged adults with CP does not show the same trend,^3,15^ though larger sample sizes are needed in older adulthood to adequately evaluate age effects. Only Ryan et al.^15^ investigated if gender was a risk factor in their medical chart review of self-disclosed falls and found that females were about 1.3 times more likely to report a fall than males. Other comorbidities or symptoms of CP have been suggested to cause falls but have not empirically studied (e.g., balance impairment, cortical visual impairment, other visual impairments, seizures, dystonia, autism, cervical stenosis, physical fatigue, mental fatigue, weakness).^10^

Larger, more robust studies are needed to inform an evidence-based fall risk stratification tool for the CP population. Real-world data on falls are currently not available for this population to build such a tool. However, fall risk is assessed routinely in healthcare settings in the United States since it is a Joint Commission requirement.^16^ These fall risk assessments provide one systematic estimate of fall risk that can be compared across patient subgroups. At our institution, the Morse Fall Scale (MFS)^17^ is administered to adult patients at outpatient visits. Our objective was to determine if fall risk, proportion of fallers, and fall-related injuries differ in adults with CP by four patient factors: GMFCS level, type of CP, age, and gender.

## Materials and Methods

The Institutional Review Board of the University of Minnesota-Twin Cities waived consent for this cross-sectional, retrospective medical chart review study. The inclusion criteria were adults (age 18 years or older) diagnosed with CP who had an in-person or virtual (telehealth) clinic visit with a physician or advanced practice provider (e.g., physiatry, infection/wound care, neurology, urology, neurosurgery, orthopedic surgery) from December 2022 to May 2023 at Gillette Children’s Phalen Adult Clinic, an outpatient clinic that provides ongoing specialty care for individuals with childhood onset disabilities. The exclusion criterion was opting out of medical records being used for research.

A CP diagnosis was determined based on the presence of an ICD-10 code from the G80 family and CP included in the Problem list of the electronic medical record. The type of CP was determined by the specific G80 code. If more than one was listed, the most descriptive G80 code was used for statistical analyses such that categories were mutually exclusive (e.g., if both other and unspecified were listed, other was chosen; if spastic diplegia and other were listed, spastic diplegia was chosen; if spastic diplegia and spastic quadriplegia were listed, spastic quadriplegia was chosen). GMFCS level was extracted from clinic visit notes or physical therapy notes. If GMFCS level was not provided in the indexed clinic visit note, then the following was used to determine GMFCS level: another clinic visit note <12 months prior, physical therapy note <12 months prior, clinic visit note >1 and up to 5 years prior, physical therapy note >1 and up to 5 years prior, and, lastly, physician co-author (LEK) assigned GMFCS level based on review of relevant information in the medical record.

Nurses or medical assistants complete the MFS for all in-person or virtual outpatient visits from self-reported information and chart review. The MFS is a 6-item questionnaire for adults to assess the likelihood of the patient falling during an inpatient stay.^17^ The six items include: history of falling in the past three months, presence of a secondary medical diagnosis, ambulatory aid type, current use of intravenous apparatus or heparin lock, gait and transferring ability, and mental status (oriented to own ability or forgets limitations). Scores range from 0 to 125.

The four study outcomes included the MFS score, percentage of individuals classified as high risk for falls based on the MFS, percentage of individuals with a history of a fall(s) in the past three months (first MFS item), and fall-related injuries. Though various cutoffs have been proposed for “high risk” of falls,^18^ the following is common and shows reasonable classification performance: low risk (0-44) and high risk (45-125). Fall-related injuries were not a required question to ask or document but were rather determined by reviewing provider’s notes. Fall circumstances were also described, if available.

For individuals who had more than one visit from December 2022 to May 2023, we selected the visit with the most outcomes available; if equivalent, we chose a random visit.

### Statistical Analysis

All analyses were performed in MATLAB using the statistical toolbox (version 2024b, MathWorks, Natick, MA). Our continuous outcome (MFS) was assessed for normality using the Shapiro-Wilk test. Non-parametric Kruskal-Wallis ANOVA was used to test the main effects of GMFCS level, type of CP, and gender. Significant pairwise comparisons were determined using Tukey’s HSD multiple comparison test. A linear regression was used to test for the main effect of age. Chi-square tests were used to assess the main effect of GMFCS level, type of CP, and gender for the two categorical outcomes: proportion of individuals classified as high risk of falls according to MFS, and the proportion of individuals who reported a fall in the past three months. A logistic regression was used to determine the effect of age for these two outcomes. Significance was assumed at p<0.05. Because of some unexpected MFS scores (e.g., 0 points), we did post hoc analyses to explore individual MFS item responses by GMFCS level.

## Results

Out of 1179 clinic visits that met the inclusion criteria over the six-month period, there were 704 unique patients, with 647 unique patients having one or more outcome available that comprised our final sample (among the 57 patient visits excluded (16 were virtual), 28 only documented that fall education was provided, 12 provider notes said no recent falls occurred, 8 cited high risk for falls). Participants ranged in age from 18-79 years and included all GMFCS levels and CP diagnoses, while non-ambulatory individuals with quadriplegia represented the vast majority (table 1). Outcomes were available for 647 participants for history of a fall(s) in the past three months and 488/647 participants (75%) for MFS score (table 2).

**Table 1.** Demographics.

|  |  |
| --- | --- |
| **n** | 647 |
| **Age (y)**, median (25%tile-75%tile), [min-max] | 32 (26-40) [18-79] |
| **GMFCS level**, n (%) |  |
| I | 25 (4%) |
| II | 92 (14%) |
| III | 75 (12%) |
| IV | 220 (34%) |
| V | 235 (36%) |
| **Type of CP**, n (%) |  |
| Spastic quadriplegic | 258 (40%) |
| Spastic diplegic | 96 (15%) |
| Spastic hemiplegic | 31 (5%) |
| Athetoid | 12 (2%) |
| Other | 151 (23%) |
| Unspecified | 99 (15%) |
| **Gender**, n (%) |  |
| Female | 289 (45%) |
| Male | 358 (55%) |

**Table 2.** Fall outcomes for the entire cohort and by GMFCS level, type of CP, and gender.

|  | **MFS score**<br>**sample**<br>**size, n** | **MFS score, median**<br>**(25%tile-75%tile),**<br>**[min-max]** | **MFS High Risk,**<br>**n (% ± standard**<br>**error)** | **Fall(s) in**<br>**prior 3**<br>**months**<br>**sample**<br>**size, n** | **Fall(s) in prior**<br>**3 months, n (%**<br>**± standard**<br>**error)** |
| --- | --- | --- | --- | --- | --- |
| **All** | 487 | 30 (15-50) [0-75] | 161 (29 ± 2%) | 647 | 44 (6 ± 1%) |
| **GMFCS level** |  |  |  |  |  |
| **I** | 17 | 35 (23-50) [15-75] | 6 (35 ± 2%) | 25 | 5 (20 ± 8%) |
| **II** | 72 | 43 (28-50) [15-75] | 32 (44 ± 2%) | 92 | 20 (22 ± 4%) |
| **III** | 48 | 50 (20-50) [15-75] | 24 (50 ± 2%) | 75 | 12 (16 ± 4%) |
| **IV** | 171 | 30 (15-40) [15-75] | 40 (23 ± 2%) | 220 | 2 (1 ± 1%) |
| **V** | 179 | 30 (15-35) [0-75] | 39 (22 ± 2%) | 235 | 2 (1 ± 1%) |
| **Type of CP** |  |  |  |  |  |
| **Spastic**<br>**quadriplegic** | 195 | 30 (15-50) [0-75] | 50 (26 ± 2%) | 258 | 4 (2 ± 1%) |
| **Spastic**<br>**diplegic** | 60 | 35 (15-50) [15-75] | 27 (45 ± 2%) | 96 | 14 (15 ± 4%) |
| **Spastic**<br>**hemiplegic** | 23 | 35 (18-50) [15-60] | 9 (39 ± 2%) | 31 | 4 (13 ± 6%) |
| **Athetoid** | 9 | 30 (15-35) [15-50] | 1 (11 ± 1%) | 12 | 0 (0 ± 0%) |
| **Other** | 121 | 30 (15-35) [15-75] | 23 (19 ± 2%) | 151 | 11 (7 ± 2%) |
| **Unspecified** | 79 | 35 (19-50) [15-75] | 31 (39 ± 2%) | 99 | 8 (8 ± 3%) |
| **Gender** |  |  |  |  |  |
| **Female** | 229 | 30 (15-50) [15-75] | 62 (27 ± 2%) | 289 | 25 (9 ± 2%) |
| **Male** | 258 | 30 (15-50) [0-75] | 79 (31 ± 2%) | 358 | 16 (4 ± 1%) |

The MFS score for the entire cohort and the subgroups of GMFCS level, type of CP, and gender are shown in table 2. For the entire cohort, the median MFS score was 30, with ranges from 0 to 75. There was a main effect for GMFCS level (Χ^2^=36.4, p<0.001). Pairwise comparisons revealed that individuals in GMFCS level II and III had higher MFS scores than those in GMFCS level IV and V (all p<0.05). There was a main effect for type of CP (Χ^2^=14.2, p=0.014). Pairwise comparisons revealed non-significant trends for higher MFS scores in individuals with unspecified CP than spastic quadriplegia (p=0.095) or other CP (p=0.067). MFS marginally increased with age (b=0.122, se=0.062, t-value=2.0, p=0.050). There was no difference by gender (Χ^2^=2.7, p=0.103).

Item-specific responses on the MFS were explored (table 3) and revealed the presence of some incorrect or ambiguous scoring. Nine individuals (2%) were documented as having no *secondary diagnosis* despite having CP. Only 35% of individuals in GMFCS level III were scored as using *crutches/cane/walker* to move which contributes 15 points, whereas the remaining were scored a 0 [wheelchair use *or* no device use]. Relatedly, two individuals in GMFCS level V were noted as using *crutches/cane/walker*, suggesting misclassification. Lastly, gait was scored as normal/bedrest/immobile for 33%, 32%, and 35% of ambulatory individuals in GMFCS levels I, II, and III, respectively. At the same time, only 75% of those in GMFCS level V were noted as bedrest/immobile/normal.

**Table 3.** Post hoc analysis of individual Morse Fall Scale items by GMFCS level.

|  | I | II | III | IV | V |
| --- | --- | --- | --- | --- | --- |
| **Secondary diagnosis, n (%)** |  |  |  |  |  |
| **No** | 0 | 2 (3%) | 2 (4%) | 2 (1%) | 3 (2%) |
| **Yes** | 18 (100%) | 73 (97%) | 46 (96%) | 180 (99%) | 190 (98%) |
| **Ambulatory aid, n (%)** |  |  |  |  |  |
| **None/bedrest/wheelchair/**<br>**nurse assist** | 18 (100%) | 64 (86%) | 31 (65%) | 175 (96%) | 191 (99%) |
| **Crutches/cane/ walker** | 0 | 10 (14%) | 17 (35%) | 7 (4%) | 2 (1%) |
| **Furniture** | 0 | 0 | 0 | 1 (1%) | 0 |
| **IV/Heparin lock, n (%)** |  |  |  |  |  |
| **No** | 17 (100%) | 74 (100%) | 48 (100%) | 182<br>(100%) | 191 (99%) |
| **Yes** | 0 | 0 | 0 | 0 | 2 (1%) |
| **Gait Impairment/Transfer ability, n (%)** |  |  |  |  |  |
| **Normal/bedrest/immobile** | 6 (33%) | 23 (32%) | 17 (35%) | 125 (68%) | 145 (75%) |
| **Weak** | 3 (17%) | 5 (7%) | 2 (4%) | 5 (3%) | 1 (1%) |
| **Impaired** | 9 (50%) | 45 (62%) | 29 (60%) | 53 (29%) | 47 (24%) |
| **Mental status, n (%)** |  |  |  |  |  |
| **Oriented to own ability** | 14 (78%) | 53 (73%) | 36 (75%) | 99 (57%) | 59 (33%) |
| **Forgets limitations** | 4 (22%) | 20 (27%) | 12 (25%) | 74 (43%) | 120 (67%) |

The percentage of individuals at high risk of falls is shown in table 2 for the entire cohort and subgroups by GMFCS level, type of CP, and gender. For the entire cohort, the percentage (standard error) at high risk of falls was 30.1 (2.1)%. Similar to the MFS score, there was a main effect for GMFCS level (Χ^2^=26.1, p<0.001) with the same significant pairwise comparisons: more individuals in GMFCS levels II and III were at high risk of falls than those in GMFCS levels IV and V (all p<0.05). There was a main effect for type of CP (Χ^2^=20.9, p<0.001); more individuals with spastic diplegia or unspecified CP were at high risk of falls compared to other CP. There was no difference by gender (Χ^2^=0.7, p=0.389) or age (b=-0.013, se=0.008, t-value=-1.6, p=0.105).

The percentage of individuals who reported a fall in the past 3 months for the entire cohort and the subgroups by GMFCS level, type of CP, and gender are shown in table 2. For the entire cohort, the percentage (standard error) of individuals who reported falling in the past three months was 6 (1)%. There was a main effect for GMFCS level (Χ^2^=79.3, p<0.001). A larger percentage of individuals in GMFCS levels I, II, and III reported falling in the past three months than those in GMFCS levels IV and V (all p<0.05). There was a main effect for type of CP (Χ^2^=24.8, p<0.001); more individuals with spastic diplegia or other CP reported falling compared to those with spastic quadriplegia (all p<0.05). Females were twice as likely to report a fall than males (Χ^2^=4.7, p=0.030). There was no difference by age (b=-0.014, se=0.013, t-value=-1.1, p=0.289).

Fall injuries and circumstances around the injury documented in clinical notes are provided in table 4. Five injuries were reported in the past three months among individuals in GMFCS levels I-III. Two other adults (ages 40s and 20s) reported injuries in the past 12 months—fractured ribs and radius. There were many individuals for whom no specific fall injury was documented, or a fall occurred outside the prior three months, but “falls frequently” or “high risk of falls” was commonly documented in the clinic note.

**Table 4.**
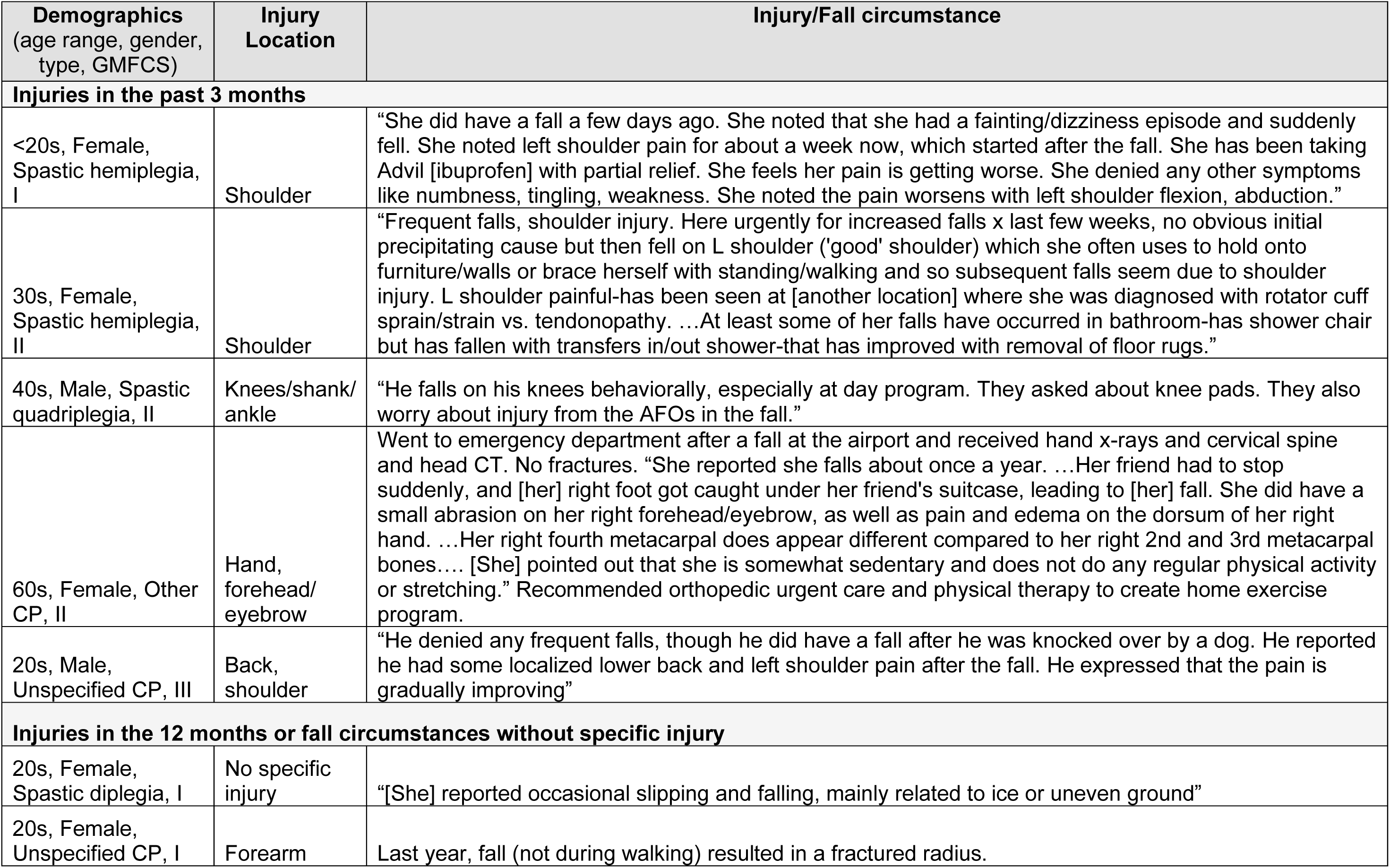
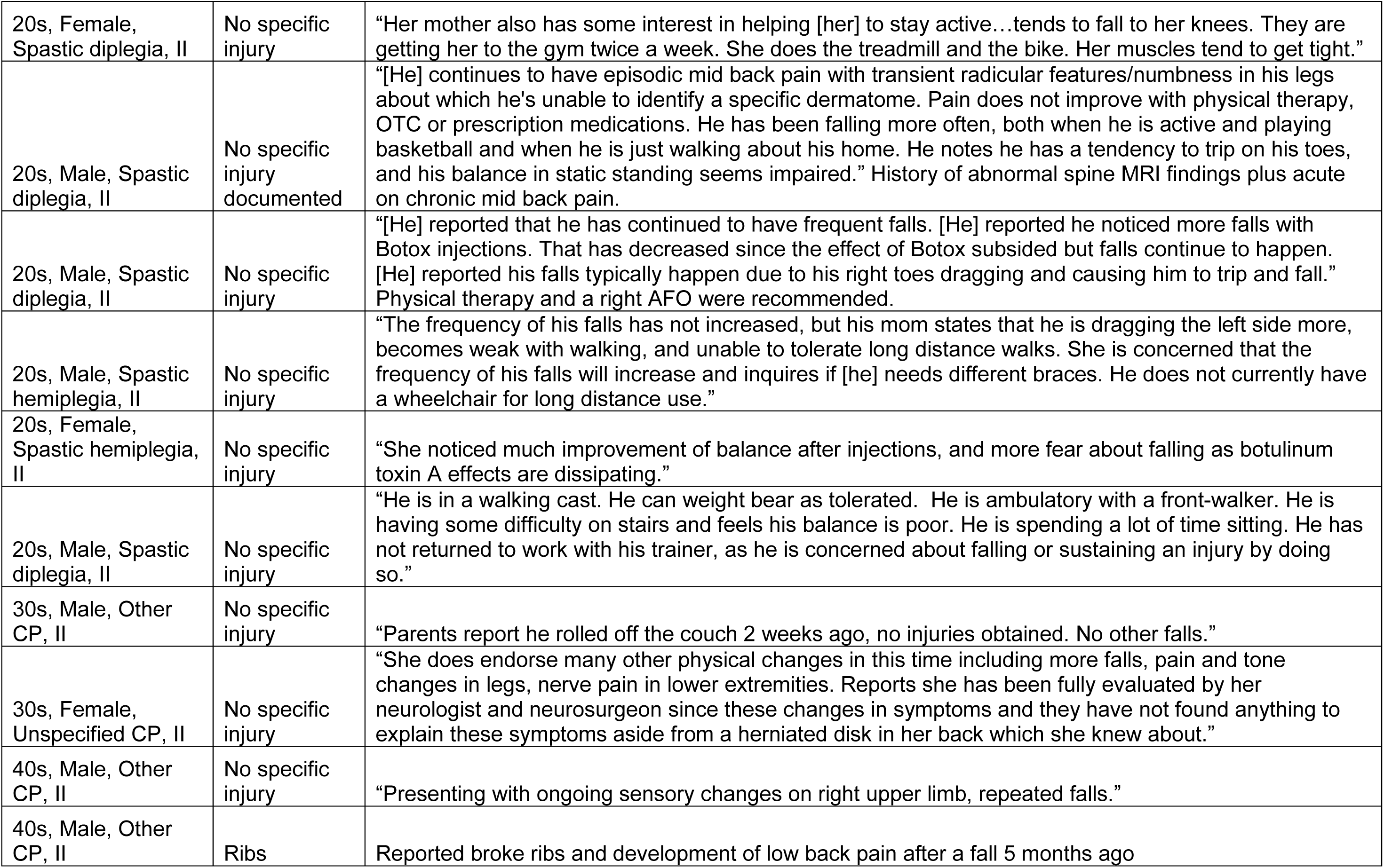
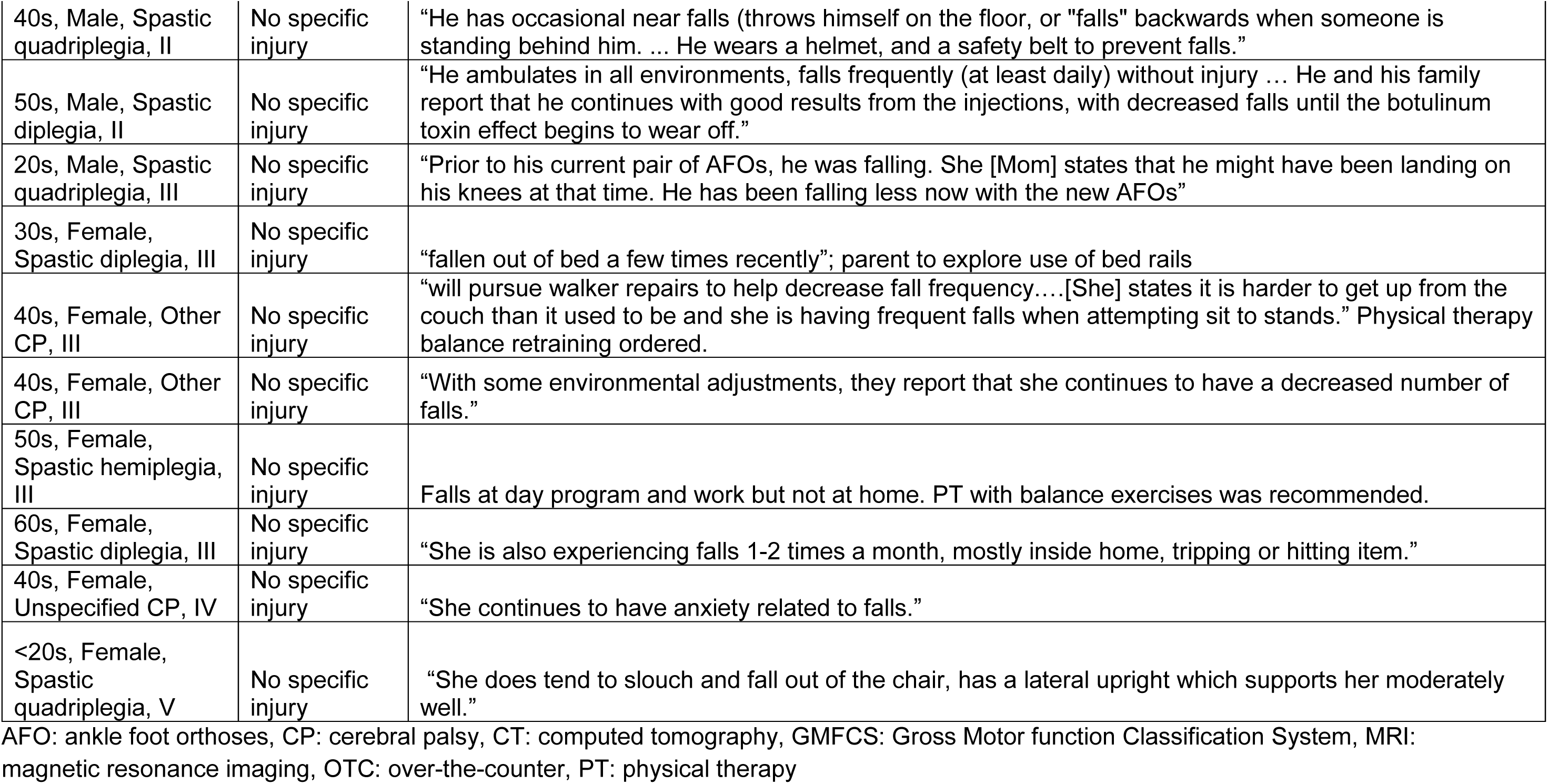
Fall injuries and/or fall circumstance.

## Discussion

The purpose of this study was to summarize falls and fall risk based on a standardized questionnaire (Morse Fall Scale) among a consecutive cohort of adults with CP seeking outpatient clinical care at a tertiary specialty clinic over a 6-month period. In our sample of 647 adult patients, ambulatory individuals had higher MFS scores, were more likely to be at high-risk for falls, and were more likely to report a fall in the past three months than non-ambulatory individuals. Females were more likely than males to report a fall. There were no or minimal differences observed for age.

Falls and fall risk differed by GMFCS level. A higher proportion of individuals in GMFCS levels I, II, and III reported a fall in the past three months compared with non-ambulatory individuals (levels IV-V), which aligns with findings in pediatric CP.^4^ Ambulatory individuals in levels II and III had higher MFS scale scores and more were categorized as high-risk compared to non-ambulatory individuals, which concurs with the pediatric and adult literature.^3,4,12^ About 15-23% of ambulatory individuals reported a fall in the past three months compared to only about 1-2% of non-ambulatory individuals. Because some individuals fall more than once per year, we cannot extrapolate these values to estimate the proportion of individuals who would fall within 12 months, though the percentages are comparable to the literature.^3,19,12,20,5,21^ Falls often occur during gait, turning, or bending among ambulatory indvidiuals,^19,3^ whereas wheelchair users often fall during transfers or reaching tasks on uneven or sloped surfaces.^22^ We hypothesize that ambulatory individuals spend more time walking or changing positions than non-ambulatory individuals spend in their respective high-risk activities, leading to a larger proportion of falls in those who can walk. Furthermore, despite just a few fall-related injuries reported and only 30% of our sample being ambulatory, all injuries were reported by ambulatory individuals. Still, severe fall-related injuries in non-ambulatory individuals do occur. Collectively, these findings suggest that falls and injuries are a more prevalent problem for ambulatory versus non-ambulatory individuals.

Falls and fall risk differed by type of CP. Individuals with spastic diplegia were more likely to report a fall than individuals with quadriplegia. To our knowledge, fall incidence and risk have not been previously compared by topography. Since individuals with spastic diplegia are more likely to be ambulatory than individuals with spastic quadriplegia and GMFCS level is predictive of falls, the observed trends match expectations. There were also differences such that individuals with unspecified CP were more likely to report a fall or be a high risk of falls than other CP. The lack of statistical difference between spastic hemiplegia and spastic diplegia is likely due to small sample sizes, though group trends suggest those with hemiplegia were less likely to report a fall or be at high risk. This underscores the importance of considering CP subtype in clinical fall-risk assessments to guide more tailored interventions or prioritize individuals facing the greatest need.

Age had no effect on high risk of falls and reported falls in the past three months though it was borderline significant for MFS score. This lack of effect/small association contradicts findings in older adults without CP where age has shown to be a strong predictor of fall risk.^13^ Studies have shown that fall incidence and balance impairment increase with age in the general population.^13^ However, the absence of an age effect agrees with the limited adult CP literature,^3,19^ though all studies were predominately young- and middle-aged adults. Prior research suggests mobility decline occurs earlier in adults with CP compared to the general population,^19,5^ and worsening balance is cited as a key contributor to their functional decline.^23^ Because people may choose to self-limit physical activity due to declining balance and function and ultimately safety concerns, they may increase sedentary time and hence be exposed to fewer opportunities to fall as they get older. This highlights the need for future studies to report falls per volume of physical activity or similar normalization method. Nonetheless, fall risk and fall incidence in young- and middle-adulthood may be driven more by CP type and GMFCS level than by chronological age.

Females were more likely to report falls than males. This is consistent with findings in adults with CP and the general population, where females report more falls, fall-related injuries, and fall-related concerns.^15,13,11,24^ Females tend to be weaker and have poorer bone health than males, predisposing them to falls and severe injuries. Alternatively, gender difference may also reflect a difference in disclosure. Sociocultural factors influence the likelihood of reporting falls and seeking medical attention, with females being more forthcoming than males.^25^ Despite this, females did not have significantly higher MFS scores, nor were they at higher risk of falls. Future research can examine how gender shapes not only the occurrence of falls and other fall risk factors but also the experiences surrounding falls and fall-related injuries. This can include gender differences in help-seeking patterns, reporting behaviors, and perceptions surrounding falls in individuals with CP.

Insufficient injury data provided in an unstandardized format precluded us from comparing injury data by patient factors. However, we noted that the seven injuries reported were all from ambulatory individuals. Both unilaterally- and bilaterally-involved individuals reported injuries. The circumstances of injurious and non-injurious falls reported revealed that most falls appeared to occur indoors and due to foot clearance or tripping. This agrees with previous findings from ambulatory individuals.^3,19^ Other behavioral, biological, and environmental causes of falls were also reported. Some notes detailed how ankle foot orthoses or botulinum toxin injections improve balance while others noted they seem to worsen balance and increase fall risk (botulinum toxin) or injury (AFO), but these case reports are not sufficient to draw conclusions. Fear of falling or concern/anxiety about falling were mentioned. Many studies have found elevated concern about falling during daily activities in adults with CP.^10,3,6,19,5,26^ A systematic investigation of fall circumstances, injuries, causes, and effects of treatment would help move the field towards fall prevention with tools such as reactive balance training, orthotic use, spasticity management, or home modifications.

### Limitations

Our study is not without limitations. As a retrospective study, one key limitation was reliance on available medical record data, i.e., the MFS, which is a fall risk screening tool designed for older adults who are inpatient with a baseline ability to ambulate. MFS score overestimation and underestimation based on item-level misclassification, which was rare for some items (secondary diagnosis) and more common for others (gait impairment), underscores the challenge of applying an inpatient-oriented measure to an outpatient heterogeneous CP population. As a result, the MFS provides only a rough estimate of fall risk. Regarding the fall history period on the MFS (three months), studies comparing recall with prospective methods demonstrate that a 3-month or 6-month recall frequently miss falls, especially falls that were non-injurious or a singular event.^27,28^ Best practice is to use a 12 month recall for a more complete fall history and to ask about number of falls to identify frequent fallers.^11^ These limitations highlight a need for a more pertinent outpatient fall risk screener or performance test(s) to assess fall risk in ambulatory and non-ambulatory people with movement disorders, or, at minimum, standardize clinical staff responses on the MFS.

Overall, there may be inaccuracy or unwillingness to disclose falls. Some clinicians at our institution believe group home staff who accompany patients may not fully know a patient’s fall history, which aligns with studies that estimate less than 40% of falls are witnessed.^29^ Unwillingness to disclose falls may stem from the negative psychosocial implications of falling, like embarrassment, fear, powerlessness, decrease self-confidence, compromised independence or autonomy, and forced dependence on mobility aids.^10,3^ Therefore, these sensitive conversations have to occur in a psychologically safe environment between clinician and patient. The World Fall Guidelines for older adults recommends clinicians routinely ask about falls rather than wait for patient disclosure, helping move towards proactive care.^11^ The guidelines also recommend swapping ‘fear of falling’ for ‘concern about falling’ to avoid negative or inaccurate connotations of ‘fear’. Lastly, the relatively small sample sizes within specific subgroups limited statistical power to detect more nuanced trends (e.g., GMFCS level I, hemiplegia, and adults older than 40 were largely underrepresented).

## Conclusion

This study identified patient factors associated with increased fall incidence and fall risk among adults with CP, namely ambulatory status (particularly GMFCS level II and III), spastic diplegia, and female gender. Adequately powered, longitudinal studies should assess both the characteristics and circumstances of falls and additional patient characteristics or comorbidities to guide targeted interventions.

## Data Availability

All data produced in the present study are available upon reasonable request to the authors

## Acknowledgments

We thank Cerebral Palsy Medical Education & Research Fund for funding Ms. Lauren Kang’s summer internship and the Endowed fund for Cerebral Palsy Treatment of Gillette Children’s for Dr. Boyer’s effort, and Rebecca Hernandez for extracting electronic medical record data.

